# Should experts guide research merit assessment? Discrepancies between Journal Impact Factor and national expert-based journal ranking list for evaluation of scientific achievement in Poland – a bibliometric analysis

**DOI:** 10.1101/2023.10.26.23297621

**Authors:** Albert Stachura, Łukasz Banaszek, Paweł K. Włodarski

## Abstract

**Objectives:** To assess discrepancies between Journal Citation Reports (JCR) Journal Impact Factor (JIF) and expert-assigned journal ranks used to evaluate achievements of medical scientists in Poland.

**Design and setting:** A bibliometric analysis based on data obtained from JCR and expert journal ranking from the Polish Ministry of Education and Science.

**Main outcome measures:** We provided descriptive analysis of all journals listed in JCR Clinical Medicine group and their respective ranks (20, 40, 70, 100, 140 or 200 points) assigned by experts in the category Medical Sciences. For each of 59 JCR Clinical Medicine categories we ranked journals from the highest JIF to the lowest (JCR category ranking) and correlated these data with points given by experts. Additionally, we analysed 4352 journals not listed in JCR but assigned expert ranks. Data collection occurred from August to September 2023.

**Results:** We extracted data on 7441 journals listed in JCR (5908 after deduplication), of which 5315 were scored by experts. Across all 6 ranks the minimum JIF was comparable and did not exceed 0.2, variances were large and outliers with JIF of 20 and above were prevalent. In 12 out of 59 JCR categories no journal was assigned 200 points. The correlation between JCR category ranking and expert ranking varied with Spearman’s r between -0.18 for Medical Informatics and -0.93 for Neuroimaging. In less than half of categories (19/59) the correlation coefficient was -0,7 or stronger. Some journals assigned 200 points in the Medical Sciences category were not related to medicine or were Polish journals with low JIF.

**Conclusions:** In Poland, the system for assessing scientific merit in medicine lacks transparency, may encourage publishing in low-quality journals and discriminate top scientists in undervalued fields. A comprehensive review of the system is necessary to promote transparent, methodologically sound, and clinically relevant research.

**What is already known on this topic:**

- Assessment of scientific merit is challenging and often based on the quality of journals, in which studies are published. Metrics like Journal Impact Factor (JIF) are surrogates of journal quality but in some countries the quality of journals is ranked by experts. Whether expert-based assessment systems promote high quality research, remains unknown.

**What this study adds:**

- Journals with JIF lower than 1 were found in all expert-assigned ranking groups and 10% of journals from Clinical Medicine JCR group were not considered in Medical Sciences Ministerial category by experts.
- Journal ranking within a study field is often not reflected by ranks assigned by experts. Scientists in every fifth JCR Clinical Medicine category cannot achieve 200 points (top expert rank) for their work despite publishing in the most prestigious journals.

**How this study might affect research, practice, or policy:**

- Current assessment system may promote publishing low-quality studies in high-rank, low-JIF journals and discourage Scientists from engaging in solving more complex but likely more relevant clinical issues.
- We advocate for a change in the assessment system, which should focus on promoting transparent, high-quality research, as well as balance discrepancies between different medical study fields.

## Introduction

An overwhelming and ever-growing amount of scientific information has created a need for evaluating scientific work, journals, and researchers. The Journal Impact Factor (JIF) is a metric based on citation count and may indicate the quality and prestige of a scientific journal (1). A study quality is often assessed based on a journal, in which it is published – hence JIF may be indirectly used to evaluate research. Such an approach isn’t flawless. Using JIF as a surrogate of a journal’s quality has been widely criticised. The San Francisco Declaration on Research Assessment in 2012 called for an end to the practice of associating JIF with scientific merit (2). In the Leiden Manifesto Hicks et al. argued the misuse of research metrics negatively affected scientific community. Authors provided ten principles of research quality evaluation, including putting more emphasis on qualitative assessment, transparency, ascertaining a high quality of locally relevant research, and accounting for variation by field of study (3).

Despite these criticisms, JIF is still used to assess scientific output (4). Many countries have created their own journal rankings to assess the research performance of local scientists and institutions. Two main approaches to creating such rankings are prevalent: (1) based solely on bibliometrics e.g., JIF or (2) determined by experts who may (or may not) take such metrics into consideration (5). The first model is used e.g., in Turkey or China, where journal ranking lists are based on data and metrics obtained exclusively from the Web of Science. The second model is used e.g., in Finland, Norway and Poland (6). Though both models rely to some degree on JIF, the latter is more subjective and likely to be shaped by the national science policy objectives. This significantly increases the risk of politicisation, which might lead to adjusting the assigned journal rank to own professional goals of experts involved in producing journal rankings, potentially creating a conflict of interests (5).

Assessment criteria have implications for funding. In Poland the funding system for scientific institutions and individual researchers is merit-based. Grants and scholarships are awarded to those publishing in top journals from the national ranking list. Rankings based on expert opinion may promote researchers, who publish in local journals of little international reputation and low JIF. In Poland, the evaluation system for scientific journals is based on points awarded by the Ministry of Education and Science (MEiN – *pol. Ministerstwo Edukacji i Nauki*) (7). The latest update of the ranking list has caused a great deal of controversy and sparked a debate in the academic community on the fairness of this quality assessment system for scientific journals. As JIF is an imperfect surrogate of journal quality, supplementing assessment systems with expert opinion may potentially help promote good research.

Here we aimed to compare the MEiN ranking system with JIF and discuss the consequences of potential discrepancies between the two models.

## Methods

### Data extraction and coding

We accessed Clarivate’s Journal Citation Reports (JCR) and downloaded bibliometric data on 7441 journals from 59 categories in the Clinical Medicine group. 2022 Journal Impact Factor for each journal was extracted. If there was no information on the current JIF or it was marked as ‘<0.1’ we assigned a value of 0. Then we accessed journal ranking list prepared by the Polish Ministry of Education and Science (MEiN) and selected 9681 titles, which were included in the ‘Medical sciences’ category. MEiN points (6 ranks: 20, 40, 70, 100, 140 and 200) were extracted for each journal. Then 2022 JIF was matched with the MEiN ranking system by the ISSN or e-ISSN number for each journal. ISSN numbers were crossed-checked against e-ISSN numbers and *vice versa* to ascertain appropriate identification and matching. If a title appeared on only one list, we assigned a value of 0 on the alternative ranking system list. We implemented an additional ranking system to sort journals within specific study fields. Within each JCR Clinical Medicine category journals were ranked from 1 to *n* (number of journals within a category) based on their JIF. Therefore, a journal with the highest JIF was assigned a rank of 1, 2^nd^ best a rank of 2 and so on. We will refer to this as to the JCR category ranking.

### Statistical analysis

All analyses were performed using R version 4.2.3. Summary statistics relating to journals included within each MEiN rank were presented. If a given journal was assigned to more than one JCR Clinical Medicine Category, it was considered only once in the summary, hence the total number of journals is smaller than 7441. For each JCR category the number of journals assigned to each MEiN rank were shown. Again, each journal was considered only once for each category. Additionally, a Spearman’s correlation coefficient was calculated to elicit associations between category ranking and MEiN ranking. We created a figure to visualise associations between Impact Factor and MEiN ranking, though included only journals with JIF below 40 as all journals with higher JIF were assigned 200 MEiN points by experts. Lower whiskers represent the minimal values, whereas the upper whiskers represent the value of Q3 + 1.5*IQR (interquartile range).

## Results

A total of 5315 journals appeared both in JCR and on the MEiN ranking list. Additionally, 593 were considered in JCR but not included under the Medical Sciences category on the MEiN list. Descriptive statistics on bibliometrics of all these journals are presented in Table 1.

**Table 1.** Descriptive statistics of journals present both in JCR and on the MEiN rakning list by MEiN ranks.

| MEiN points | Number of journals | Min IF | Q1 | Median JIF | Mean JIF | Q3 | Max JIF |
| --- | --- | --- | --- | --- | --- | --- | --- |
| 0 | 593 | 0 | 1 | 1.8 | 2.26 | 2.9 | 17.6 |
| 20 | 1242 | 0 | 0.3 | 0.9 | 1.68 | 2 | 38.2 |
| 40 | 939 | 0 | 1 | 1.5 | 1.87 | 2.2 | 28.1 |
| 70 | 1325 | 0 | 1.8 | 2.5 | 2.75 | 3.2 | 36.4 |
| 100 | 1071 | 0 | 2.7 | 3.4 | 3.84 | 4.6 | 24.8 |
| 140 | 541 | 0.2 | 3.8 | 5.3 | 6.15 | 7.5 | 37.3 |
| 200 | 197 | 0.2 | 9 | 13.9 | 24.34 | 28.4 | 254.7 |

JIF – Journal Impact Factor, MEiN – Ministry of Education and Science (*pol. Ministerstwo Edukacji i Nauki*), Q1 – 1<sup>st</sup> quartile, Q3 – 3<sup>rd</sup> quartile

A total of 4352 journals were assigned MEiN rank but were not listed in JCR. Of them, 2716 (62,4%) were assigned 20 points, 617 (14,2%) - 40 points, 472 (10,8%) - 70 points, 303 (7%) - 100 points, 162 (3,7%) - 140 points and 82 (1,9%) - 200 points.

JIF of all journals with Impact Factor lower than 40 was plotted against MEiN ranking and presented in Figure 1. The mean JIF grows successively in journals assigned between 20 and 140 MEiN points and is the highest among those assigned 200 points. The mean JIF in the group of journals with assigned JIF and not assigned MEiN points was higher than in the group with 20 or 40 MEiN points. In all groups there were numerous outliers, some of them with JIF over 20 despite having been assigned between 20 and 140 MEiN points. At the same time, minimal JIF values for all groups were comparable even among those assigned 200 MEiN points. The largest JIF variance is also seen in this group.

**Figure 1.**
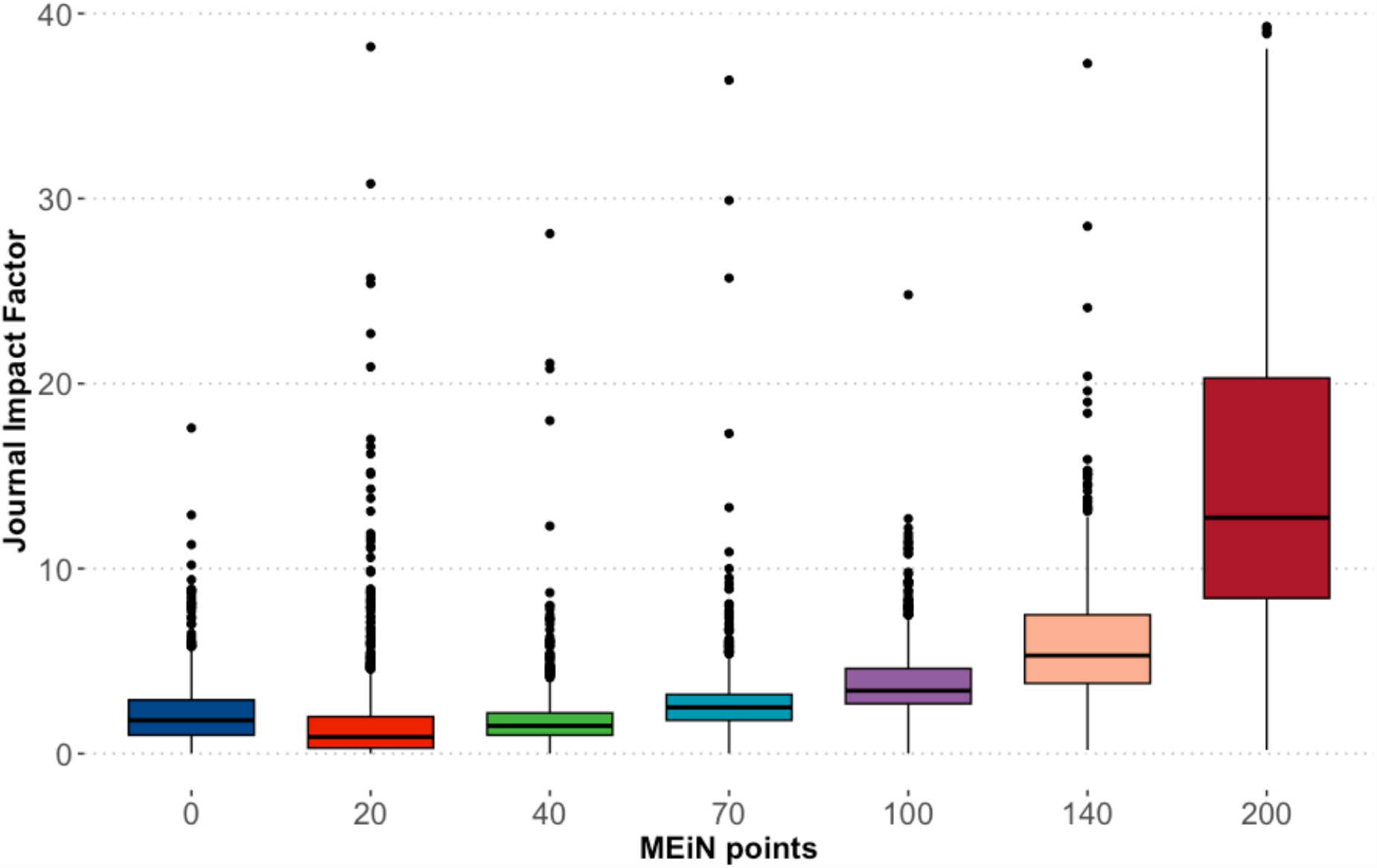
Relationship between Journal Impact Factor and MEiN points by MEiN ranks for journals with JIF below 40. Lower whiskers represent the minimal values, whereas the upper whiskers represent the value of Q3 (3^rd^ quartile) + 1.5*IQR (interquartile range).

The numbers of journals assigned different MEiN ranks and included in 59 JCR Clinical Medicine categories are shown in Table 2. For twelve categories (Andrology, Audiology & speech-language pathology, Dermatology, Integrative & complementary medicine, Medical informatics, Medical laboratory technology, Medicine, legal, Nursing, Otorhinolaryngology, Rehabilitation, Substance abuse and Tropical Medicine) no journal was assigned 200 MEiN points. The correlation between JCR Category ranking and MEiN ranking varied considerably. The lowest was noted for Medical Informatics (r = -0,18) and the highest for Neuroimaging (r = -0,93). In most cases the strength of correlation was moderate.

**Table 2.** Distribution of journals assigned different MEiN ranks by JCR Clinical Medicine categories.

| JCR Category | MEiN points |  |  |  |  |  |  | Max JIF | r |
| --- | --- | --- | --- | --- | --- | --- | --- | --- | --- |
|  | 0 | 20 | 40 | 70 | 100 | 140 | 200 |  |  |
| Allergy | 0 | 8 | 6 | 6 | 10 | 7 | 1 | 14.2 | -0,78 |
| Andrology | 0 | 1 | 1 | 5 | 0 | 1 | 0 | 4.8 | -0,31 |
| Anesthesiology | 0 | 18 | 14 | 12 | 13 | 5 | 1 | 10.7 | -0,85 |
| Audiology & speech-language pathology | 4 | 3 | 1 | 10 | 13 | 4 | 0 | 3.7 | -0,58 |
| Behavioral sciences | 20 | 0 | 1 | 13 | 14 | 3 | 3 | 29.3 | -0,62 |
| Cardiac & cardiovascular systems | 1 | 52 | 49 | 46 | 33 | 18 | 12 | 49.6 | -0,72 |
| Clinical neurology | 2 | 44 | 42 | 62 | 75 | 32 | 11 | 48 | -0,68 |
| Critical care medicine | 1 | 15 | 4 | 10 | 11 | 6 | 5 | 76.2 | -0,78 |
| Dentistry, oral surgery & medicine | 2 | 46 | 27 | 27 | 37 | 16 | 2 | 18.6 | -0,75 |
| Dermatology | 3 | 20 | 16 | 29 | 17 | 8 | **0** | **13.8** | -0,70 |
| Emergency medicine | 0 | 21 | 11 | 12 | 6 | 3 | 1 | 8 | -0,64 |
| Endocrinology & metabolism | 1 | 28 | 16 | 49 | 58 | 20 | 11 | 44.5 | -0,74 |
| Engineering, biomedical | 30 | 9 | 20 | 22 | 17 | 13 | 5 | 28.1 | -0,28 |
| Gastroenterology & hepatology | 2 | 32 | 21 | 29 | 36 | 10 | 9 | 65.1 | -0,72 |
| Genetics & heredity | 10 | 27 | 19 | 48 | 42 | 30 | 14 | 42.7 | -0,63 |
| Geriatrics & gerontology | 4 | 13 | 11 | 13 | 19 | 9 | 1 | 16.6 | -0,59 |
| Health care sciences & services | 44 | 22 | 15 | 36 | 31 | 16 | 2 | 20.9 | -0,45 |
| Health policy & services | 33 | 8 | 20 | 23 | 23 | 7 | 2 | 11.5 | -0,35 |
| Hematology | 0 | 18 | 16 | 22 | 23 | 13 | 4 | 28.5 | -0,70 |
| Immunology | 4 | 16 | 11 | 49 | 50 | 32 | 15 | 100.3 | -0,69 |
| Infectious diseases | 1 | 19 | 15 | 38 | 36 | 14 | 5 | 56.3 | -0,58 |
| Integrative & complementary medicine | 5 | 10 | 10 | 11 | 5 | 2 | **0** | **7.9** | -0,63 |
| Materials science, biomaterials | 20 | 2 | 4 | 10 | 7 | 10 | 2 | 18.9 | -0,38 |
| Medical ethics | 6 | 4 | 3 | 1 | 5 | 2 | 2 | 13.4 | -0,37 |
| Medical informatics | 11 | 10 | 1 | 8 | 9 | 3 | **0** | **30.8** | -0,18 |
| Medical laboratory technology | 3 | 5 | 5 | 11 | 4 | 4 | **0** | **10** | -0,75 |
| Medicine, general & internal | 8 | 155 | 64 | 52 | 22 | 11 | 13 | 168.9 | -0,73 |
| Medicine, legal | 2 | 4 | 4 | 6 | 6 | 2 | **0** | **3.4** | -0,84 |
| Medicine, research & experimental | 5 | 46 | 27 | 39 | 45 | 19 | 8 | 82.9 | -0,62 |
| Neuroimaging | 0 | 1 | 1 | 5 | 6 | 1 | 1 | 5.7 | -0,93 |
| Neurosciences | 7 | 35 | 20 | 80 | 83 | 63 | 18 | 34.7 | -0,61 |
| Nursing | 122 | 14 | 23 | 19 | 10 | 2 | **0** | **8.1** | -0,23 |
| Nutrition & dietetics | 25 | 13 | 10 | 23 | 23 | 12 | 2 | 13.6 | -0,51 |
| Obstetrics & gynecology | 0 | 37 | 20 | 41 | 22 | 9 | 1 | 13.3 | -0,67 |
| Oncology | 1 | 65 | 32 | 83 | 72 | 48 | 17 | 254.7 | -0,58 |
| Ophthalmology | 1 | 20 | 12 | 33 | 17 | 10 | 2 | 17.8 | -0,63 |
| Orthopedics | 2 | 32 | 24 | 27 | 25 | 16 | 2 | 12.1 | -0,61 |
| Otorhinolaryngology | 2 | 13 | 9 | 18 | 17 | 4 | **0** | **7.8** | -0,76 |
| Pathology | 1 | 11 | 16 | 28 | 19 | 9 | 3 | 36.2 | -0,73 |
| Pediatrics | 4 | 43 | 44 | 46 | 34 | 12 | 2 | 36.4 | -0,64 |
| Peripheral vascular disease | 1 | 16 | 15 | 28 | 21 | 7 | 5 | 37.8 | -0,66 |
| Pharmacology & pharmacy | 57 | 44 | 48 | 97 | 67 | 40 | 9 | 120.1 | -0,62 |
| Primary health care | 0 | 6 | 8 | 7 | 5 | 0 | 1 | 6.1 | -0,33 |
| Psychiatry | 13 | 39 | 47 | 70 | 55 | 32 | 9 | 73.3 | -0,61 |
| Psychology, clinical | 76 | 8 | 29 | 27 | 22 | 14 | 2 | 18.4 | -0,46 |
| Public, environmental & occupational health | 48 | 69 | 79 | 95 | 69 | 28 | 11 | 50 | -0,41 |
| Radiology, nuclear medicine & medical imaging | 8 | 41 | 39 | 59 | 33 | 17 | 6 | 19.7 | -0,57 |
| Rehabilitation | 15 | 31 | 26 | 48 | 34 | 12 | **0** | **12.1** | -0,56 |
| Reproductive biology | 5 | 4 | 4 | 8 | 7 | 5 | 2 | 13.3 | -0,55 |
| Respiratory system | 0 | 23 | 14 | 28 | 25 | 6 | 4 | 76.2 | -0,73 |
| Rheumatology | 1 | 19 | 8 | 7 | 9 | 8 | 3 | 33.7 | -0,72 |
| Sport sciences | 31 | 15 | 13 | 19 | 26 | 12 | 5 | 18.4 | -0,56 |
| Substance abuse | 8 | 3 | 12 | 13 | 16 | 2 | 0 | 9.4 | -0,53 |
| Surgery | 2 | 75 | 55 | 60 | 62 | 23 | 7 | 16.9 | -0,78 |
| Toxicology | 7 | 9 | 19 | 32 | 21 | 11 | 2 | 12.5 | -0,55 |
| Transplantation | 0 | 7 | 5 | 7 | 8 | 4 | 1 | 8.9 | -0,72 |
| Tropical medicine | 0 | 3 | 7 | 11 | 6 | 1 | 0 | 8.1 | -0,45 |
| Urology & nephrology | 3 | 30 | 25 | 38 | 14 | 12 | 3 | 41.5 | -0,62 |
| Virology | 1 | 5 | 4 | 12 | 12 | 6 | 1 | 30.3 | -0,48 |

JCR – Journal Citation Reports, JIF – Journal Impact Factor, MEiN – Ministry of Education and Science (*pol. Ministerstwo Edukacji i Nauki*), $r$ – Spearman's correlation coefficient for associations between JCR category ranking and MEiN ranking.

## Discussion

The latest MEiN ranking list was released on the July 18^th^ 2023, almost exactly one year after the 2022 Journal Citation Report had been announced (June 2022). MEiN points are used in Poland to assess scientific achievements and are crucial for receiving grants, scholarships, and academic titles. They are also used to evaluate academic staff and individual departments influencing money distribution within scientific institutions. It could be assumed the national ranking would be relatable to the internationally recognised JIF list. Our analysis showed that these two metrics do not always agree and frankly are sometimes at odds with each other.

We found that about 10% of journals included in JCR Clinical Medicine group has not been considered by MEiN within its Medical Sciences category. Some of the left-out titles had a JIF of over 17. Across ranks, minimal JIFs were virtually the same (around 0) and variations of JIF values were considerable. In extreme cases, journals with Impact Factor of over 30 were assigned 20 MEiN points, while those with JIF of 0,2 were included in the 200 MEiN points group. The number of journals included within each subsequent rank was not decreasing, as one would expect, but was irregular. More journals were assigned 70 or 100 points as compared to only 40. Ranks 140 and 200 were the most elite comprising a total of 738 journals. When we analysed MEiN ranks distribution across JCR categories, we found that every fifth category lacked a single journal with a rank of 200 points. It means publishing an article awarded 200 points in fields such as andrology, dermatology, or otorhinolaryngology is impossible. Strikingly, some of the best journals included in JCR categories, not assigned a rank of 200, had JIF of between 3.4 and 30.8. Moreover, in many cases JCR category ranking correlated moderately with MEiN ranking. It suggests experts scoring was informed by more than just JIF-based prestige of a given journal within a field. What guided their decision remains unanswered.

It seems clear that some of the most prestigious journals were strongly undervalued by the experts assigning MEiN ranks, as explained above. What about the journals considered worthy of 200 MEiN points? Among these are familiar titles – *The New England Journal of Medicine, The Lancet, JAMA, The BMJ, Annals of Internal Medicine* and so on. However, some lesser-known titles were also included in this category - *Medycyna Pracy* (Polish journal of occupational medicine), *Archives of Budo, Acta Angiologica* with JIF of 0.2. Some titles not listed in the Clinical Medicine group by JCR were also assigned 200 points – *The Journal of Agricultural Science, Journal of Antropological Archeology, Soil Biology and Biochemsitry, Disaster and Emergency Medicine Journal* (not yet assigned JIF). When applying for grants or scholarships, Budo enthusiasts or scientists publishing in local journals are assessed alongside researchers publishing in the top medical journals. All these groups will have equal chances, which is clearly unfair and may discourage scientists from engaging in time-consuming and complex but (likely) more meaningful work.

Impact Factor was first introduced by Eugene Garfield in 1975 and was meant to become an indicator of usage of scholarly literature, as well as help identify potential venues for publication, especially for interdisciplinary research (8). It has been deemed a valid measure of journal quality among researchers and practicing physicians (9) but also received criticism as a bibliometric indicator (8). JIF opponents argue that its calculation is prone to asymmetry between numerator and denominator, journal self-citations are included, length of citation window is inaccurate, citation distribution is skewed, some disciplines are undervalued, and JIF gets inflated over time. But for all these criticisms, JIF remains a widely used bibliometric indicator and introduction of any new metric should mitigate with rather than replicate its flaws. MEiN system lacks a transparent calculation system, encourages publishing in high-rank low-quality journals (contributing to self-citation) and failed to address discrepancies between medical disciplines, as shown above.

Any ranking system should ideally aim to promote researchers doing ‘good’ research. What constitutes a good quality medical research has been widely discussed and studied. In his famous editorial, Doug Altman argued that ‘*We need less research, better research, and research done for the right reasons*’ and ‘(…) *much poor research arises because researchers feel compelled for career reasons to carry out research that they are ill equipped to perform, and nobody stops them*’ (10). Altman advocated for research transparency and was the founder of the EQUATOR (Enhancing the QUAlity and Transparency Of health Research) Network, promoting good reporting practices (11). As proven later, better reporting quality is associated with a lower risk of bias (and hence better quality) in randomised controlled trials (12) and it seems reasonable to assume it may be true for other study designs as well.

The Cochrane Methodology Review Group created a list of outcome measures that would facilitate identifying a good quality biomedical study. It ideally should be important, useful, relevant, methodologically sound, ethical, complete and accurate (13). A gold standard editorial process should ascertain all these qualities in published research. Higher JIF is associated with better adherence to reporting guidelines in health care literature (14) – a surrogate outcome for methodological soundness (13). We do not argue a higher JIF must equal better research quality. However, a process of publishing in a high impact journal requires an iterative process to improve one’s work, likely producing a better quality and more informative study. Does the current MEiN ranking system encourage publishing good quality articles in journals advocating for transparency of reporting? Likely not.

The process by which the current MEiN ranking list was created lacks transparency. No clear criteria for assigning ranks were published. Much like JIF, MEiN ranking helps identify venues for publication – particularly journals ranked high, which are relatively easy to publish in. The key question comes to mind: is it policymakers’ job to artificially inflate the value of local journals and even put them on a par with top medical titles? In our view, more emphasis should be put on improving the quality of peer-review, editorial process, and strict adherence to reporting guidelines to promote transparent and reproducible research. Instead of encouraging scientists to publish poor studies in ‘high-rank’ journals, they should be equipped with skills Altman argued few possessed. Courses in basic medical statistics and statistical inference, research design and critical appraisal should be promoted. The joined effort of editors and researchers would likely result in better research addressing significant clinical problems to find better solutions for patients.

Our analysis has some limitations. We only extracted data on journals included in the Medical Sciences category by MEiN. Therefore, some of the titles from JCR Clinical Medicine list that are not present within this MEiN category could be present in different ones. We assumed that clinical medicine is a narrower term than medical sciences and that the latter should contain all the journals from the former group. As shown above, some journals unrelated to health care sciences were also included by MEiN in the Medical Sciences group. How each title was assigned to given categories remains unclear. Another limitation is that authors of this study are Polish scientists who undergo evaluation based on the said ranking and therefore may be biased. However, we aimed to analyse bibliometric data in an objective, quantitative way and focused on the most extreme outliers, regardless of our own primary field of study.

### Recommendations

It is debatable whether expert opinion adds value to the metric-based assessment systems. On the one hand such insight may help balance discrepancies between study disciplines. Experts can help identify journals with strict editorial criteria publishing good quality research – something a metric based on citation count may fail to do. On the other hand, involving experts in the assessment process creates a risk of conflicts of interests and hindering transparency of journal rank assignment. We do not argue any system is superior over another. However, based on our analysis we share recommendations that could help improve expert-based journal ranking lists.

- A clear and transparent method of rank assignment (or points calculation) should be provided in updated versions of MEiN ranking list.
- Strict criteria for assigning top ranks should be applicable across all disciplines. These could include minimal JIF and journal’s transparency of reporting requirements.
- Journals ranked 100 points or higher should all have JIF.
- Journals with JIF not included in the current version should be evaluated and considered in future ranking lists.
- Experts from all fields should be included in the ranking list creation process to avoid undervaluing certain disciplines. Moreover, all people involved should claim their conflicts of interests, particularly editorial functions.
- Type of study design should be considered when assessing scientific merit. Randomised controlled trials or meta-analyses should not be compared with case reports as the latter require way less effort.

## Conclusion

In Poland, medical research assessment is based on a non-transparent and unbalanced system created by experts. It may encourage publishing low-quality research in journals assigned high expert rank but with low JIF. A comprehensive review of the journal ranking list is needed to promote researchers adequately addressing relevant clinical problems. It is our hope our analysis will spark discussion in other countries currently using similar expert-based assessment systems.

## Data Availability

Data are available upon reasonable request from Albert Stachura. That includes the lists of journals with their JIF and matched MEiN points.

## Acknowledgments

We thank Wiktor Paskal, an associate professor from the Department of Methodology, Medical University of Warsaw, for his research assistance during the project.

## Contributors

AS conceived the general thesis and is the guarantor of the study. AS and ŁB extracted and analysed data, wrote the initial draft, and edited the manuscript. PW edited the manuscript and helped with data interpretation. All authors verified and approved of the final version of the manuscript.

## Patient and public involvement

Patients and/or the public were not involved in the design, conduct, reporting, or dissemination plans of this research.

## Patient consent for publication

Not applicable.

## Ethics approval

Not applicable

## Provenance and peer review

Not commissioned; externally peer reviewed.

## References

1. Garfield E. The history and meaning of the journal impact factor. JAMA. 2006;295(1):90–3.

2. Cagan R. The San Francisco Declaration on Research Assessment. Dis Model Mech. 2013;6(4):869–70.

3. Hicks D, Wouters P, Waltman L, de Rijcke S, Rafols I. Bibliometrics: The Leiden Manifesto for research metrics. Nature. 2015;520(7548):429–31.

4. Smart P. Is the impact factor the only game in town? Ann R Coll Surg Engl. 2015;97(6):405–8.

5. Lowry PB, Gaskin J, Humpherys SL, Moody GD, Galletta DF, Barlow JB, et al. Evaluating journal quality and the association for information systems senior scholars’ journal basket via bibliometric measures: Do expert journal assessments add value? MIS quarterly. 2013:993–1012.

6. Kulczycki E, Huang Y, Zuccala AA, Engels TC, Ferrara A, Guns R, et al. Uses of the Journal Impact Factor in national journal rankings in China and Europe. Journal of the Association for Information Science and Technology. 2022;73(12):1741–54.

7. Polish Ministry of Education and Science (MEiN). Journal ranking list [online]. 2023. https://www.gov.pl/web/edukacja-i-nauka/nowy-rozszerzony-wykaz-czasopism-naukowych-i-recenzowanych-materialow-z-konferencji-miedzynarodowych (accessed 26 October 2023).

8. Lariviere V, Sugimoto CR. The journal impact factor: A brief history, critique, and discussion of adverse effects. Springer handbook of science and technology indicators. 2019:3–24.

9. Saha S, Saint S, Christakis DA. Impact factor: a valid measure of journal quality? J Med Libr Assoc. 2003;91(1):42–6.

10. Altman DG. The scandal of poor medical research. Bmj. 1994;308(6924):283–4.

11. Simera I, Moher D, Hirst A, Hoey J, Schulz KF, Altman DG. Transparent and accurate reporting increases reliability, utility, and impact of your research: reporting guidelines and the EQUATOR Network. BMC Med. 2010;8:24.

12. Tikka C, Verbeek J, Ijaz S, Hoving JL, Boschman J, Hulshof C, et al. Quality of reporting and risk of bias: a review of randomised trials in occupational health. Occup Environ Med. 2021;78(9):691–6.

13. Jefferson T, Rudin M, Brodney Folse S, Davidoff F. Editorial peer review for improving the quality of reports of biomedical studies. Cochrane Database Syst Rev. 2007;2007(2):Mr000016.

14. Samaan Z, Mbuagbaw L, Kosa D, Borg Debono V, Dillenburg R, Zhang S, et al. A systematic scoping review of adherence to reporting guidelines in health care literature. J MulEdiscip Healthc. 2013;6:169–88.

